# Agent-based modeling of DFNB1A prevalence with regard to intensity of selection pressure in isolated human population: will cochlear implantation increase the cases of hereditary deafness?

**DOI:** 10.1101/2021.08.11.21261942

**Authors:** Georgii P. Romanov, Anna A. Smirnova, Vladimir I. Zamyatin, Aleksey M. Mukhin, Fedor V. Kazantsev, Vera G. Pshennikova, Fedor M. Teryutin, Aisen V. Solovyev, Sardana A. Fedorova, Olga L. Posukh, Sergey A. Lashin, Nikolay A. Barashkov

## Abstract

It was evidenced, that the increase in the prevalence of autosomal recessive deafness 1A (DFNB1A) in populations of European descent was promoted by assortative marriages among deaf people. Assortative marriages become possible with a widespread introduction of sign language resulting in increased the genetic fitness of deaf individuals, thus relaxing selection against deafness. Currently, cochlear implantation is becoming a common method of rehabilitation for deaf patients, restoring their hearing ability and promoting the acquirement of spoken language. Whether the mass cochlear implantation could affect the spread of hereditary deafness is unknown. We have developed an agent-based computer model for analysis of the spread of DFNB1A. Using the model, we tested impact of different intensity of selection pressure on an isolated human population for 400 years. The modeling of the “purifying” selection pressure on deafness resulted in decrease of the proportion of deaf individuals and the pathogenic allele frequency. The modeling of relaxed selection resulted in increase of the proportion of deaf individuals and the decrease of the pathogenic allele frequency. The results of neutral selection pressure modeling showed no significant changes in both the proportion of deaf individuals and the pathogenic allele frequency after 400 years. Thus, initially low genetic fitness of deaf people can be significantly increased in the presence of assortative mating by deafness, resulting in a higher prevalence of DFNB1A. Contrary, frequency of pathogenic allele and the incidence of hereditary hearing loss will not increase in a population where all deaf individuals undergo cochlear implantation.

## INTRODUCTION

Hearing loss (HL), caused by both environmental and genetic factors, affects more than 10% of the world’s population, leads to disability and significantly reduces the quality of life of deaf individuals. On average, one per 1000 newborns are born deaf, and in 50-60% of cases, the pathology has a genetic cause [1, 2]. Hereditary HL cases are subdivided in non-syndromic (isolated HL) and syndromic (HL and other clinical traits) forms. Currently, more than 400 HL-associated syndromes are described, comprising ∼ 30% of all HL cases and ∼ 70% are accounted to non-syndromic HL [3]. Hereditary non-syndromic HL is a monogenic disease with unique high genetic heterogeneity. To date, about 160 genetic loci associated with non-syndromic HL are known, and about 123 genes have been identified, mutations in which lead to hearing impairment [4]. The autosomal recessive deafness 1A (DFNB1A) caused by mutations in the *GJB2* gene (MIM 121011, 13q12.11), encoding the protein connexin 26 (Cx26), is the most prevalent in many populations [5]. The proportion of DFNB1А among hereditary forms of HL is 17.3% worldwide, and reaches up to 27.1% in populations of European descent [5]. In total, about 400 mutations in the *GJB2* gene are known and the majority of them are recessively inherited [6]. Varying prevalence of different *GJB2* mutations has been shown for populations worldwide that can be explained by the high scale population history events [7-13]. The unique *GJB2* mutational spectrum and the accumulation of certain *GJB2* mutations in different ethnic groups can be attributed to founder effect [14-22].

In addition, Nance et al. (2000, 2004) suggested that the certain social factors could be a strong driving force behind the increased incidence of DFNB1A in developed countries due to relaxed selection against deafness which occurred after the introduction of sign language 400 years ago in many Western countries and the subsequent establishment of residential schools for the deaf [23, 24]. Sign language is the main type of communication of people with congenital HL. Linguistic homogamy (the ability to communicate in sign language) promoted the assortative marriages between deaf partners, improved living conditions (social, educational, and economic circumstances) of deaf individuals and led to significantly increased genetic fitness (reproductive capabilities) of them. Using computer modeling, it was showed that the joint effect of high rate of assortative marriages and relaxed selection against deafness could have doubled the frequency of DFNB1A in the United States during the past 200 years [24]. The computer modeling results were later supported by the comparative analysis of modern and retrospective demographic parameters of deaf population of USA [25, 26]. Thus, it was evidenced, both theoretically and practically, that the increase in the HL prevalence in the USA was a consequence of the increased genetic fitness of deaf individuals which was caused by linguistic homogamy among them.

Currently, the rate of assortative mating among deaf people in different countries is reaching up to 90% [26, 27], however, the increasing availability of cochlear implantation can certainly affect the mating structure of deaf people. Theoretically, deaf people with genetic forms of HL, including those caused by biallelic recessive *GJB2* mutations, who undergo cochlear implantation, acquire the ability to hear, that is, they become phenotypically hearing individuals, despite their pathogenic genotypes. Thus, it is highly possible that such individuals will be directed into hearing mating pool and, consequently, alter the prevalence of hereditary HL. Moreover, whether relaxed selection will have similar effect in a population with genetic structure different to the demonstrated one? One of potential approaches to address these issues is a computer modeling. Computer modeling provides excellent opportunities of analyzing a large scale population-genetics dynamics in real-time. Among different modeling approaches, the agent-based modeling is the most flexible way to simulate genetic data, as they allow one to simulate very specific behavior of individuals depending on the environment. However, for purposes of correct modeling of spread of DFNB1A, an isolate population with known data on prevalence of *GJB2* gene mutations causing hereditary HL, and data on proportion of assortative mating among deaf people and their reproductive capabilities in comparison with their hearing siblings is needed.

Thus, the aim of this study is a computer modeling of the DFNB1A prevalence in an isolated human population with known demographic parameters with regard to genetic fitness of deaf individuals.

## MATERIALS AND METHODS

### The computer model

To assess the prevalence of a congenital recessive HL under reduced or absent selection pressure for deafness, we developed an agent based model which simulates the population dynamics. We have used C++ programming language and Microsoft Visual Studio 2019 development environment. The key element of the model is a decentralized agent which is an individual with a given pattern of behavior depending on his phenotype and “environmental factors”. The environment for an individual agent is a whole population consisting of other agents. Agents do not have age and “live” for only one generation (1 generation = 20 years). The generations in the model are non-overlapping. In each generation (cycle), the main algorithm runs the processes of selection of marital partners for agents based on the phenotype (deaf / hearing, knowledge of sign language) and birth of children, depending on the genotypes of parents (Figure 1). The algorithm of the program is specified in the Supplementary Materials. For batch operation of the program and statistical processing of the output data, we developed a service script which controls the number of simulation runs, sets the starting parameters for the model, performs statistical calculations and generates summary plots. The script is written in the Python programming language using the `pandas` and `matplotlib` libraries.

**Figure 1.**
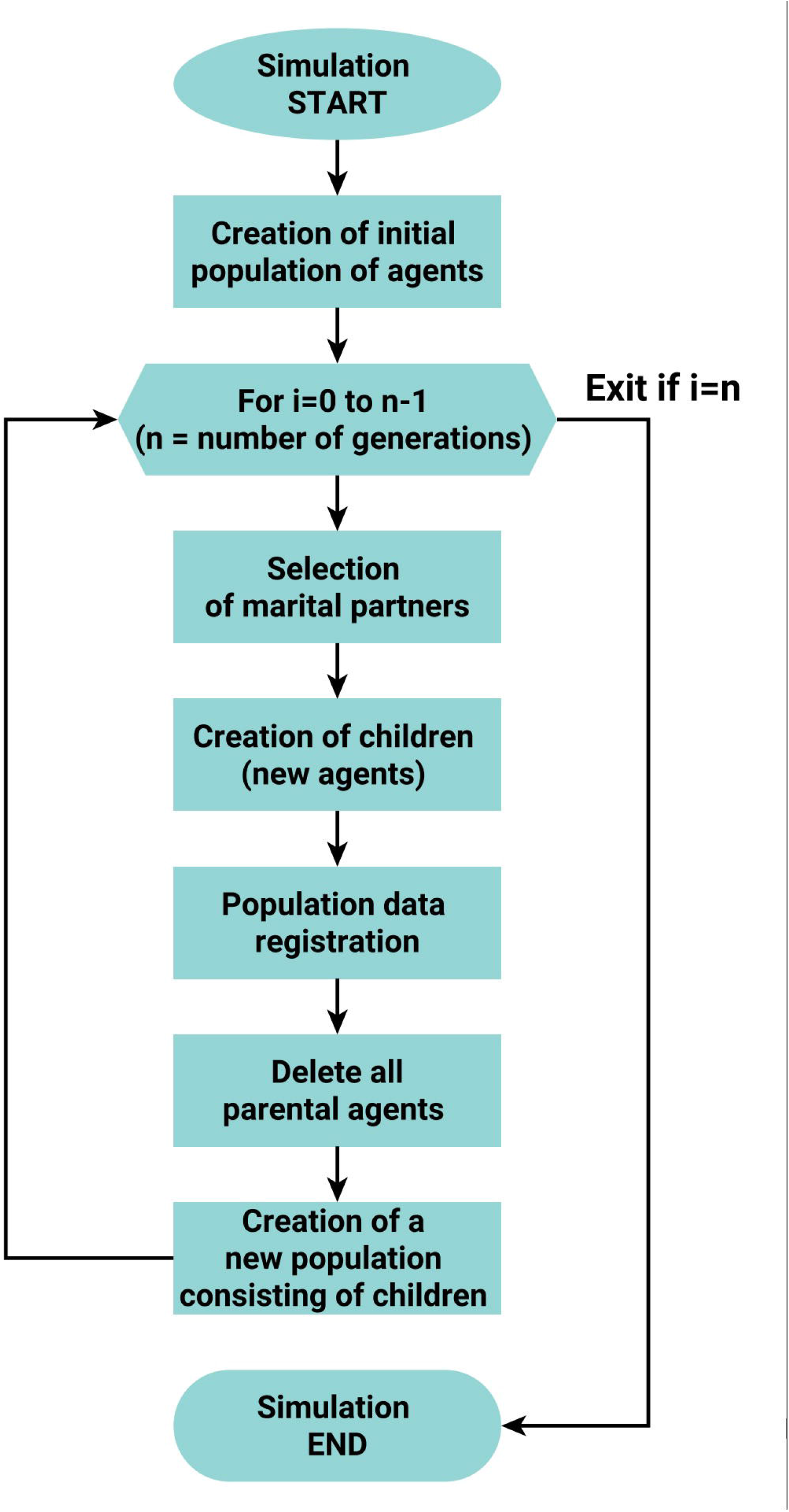
The simplified scheme of main cycle of the program. At the initial stage, a population of agents (individuals) is created according to the parameters set by the user. The population of next generation consists of the progeny of the agents of the previous generation.

We carried out the simulations with three different sets of combinations of initial parameters of the model population (Supplementary Table 1). In the first scenario, the agents (individuals) did not know sign language and there was no deaf community (parameters `sign_lang_d = 0`; `deaf_community_model = 0`). In the second scenario, deaf individuals used sign language and formed a community (`sign_lang_d = 1`; `deaf_community_model = 1`). In the third scenario, all agents were phenotypically hearing regardless of their genotype (`sign_lang_d=1`; `sign_lang_h = 1`; `deaf_community_model = 0`), representing the mass introduction of cochlear implantation in the population. The parameters for each scenario are presented in Supplementary Table 1. To generate reliable statistical data, the simulation of each scenario was performed 1,000 times. Statistical processing was carried out by calculating 99% confidence intervals for each set of values (n = 1,000) of the variables produced by the program.

### Verification of the model

To confirm the validity of the data produced by the model, we conducted additional simulations. In this scenario, the initial allele frequencies, genotypes, deaf reproductive capacity, and proportion of assortative marriages were consistent with previously published data on the US deaf population [24-26]. When simulated with these parameters, the model produced data corresponding to the dynamics of hereditary HL in the U.S. over 200 years (Supplementary Table 5, Supplementary Figure 5) [24-26].

## RESULTS

We have developed an agent-based computer model for analysis of the spread of hereditary congenital recessive HL in an isolated human population (Supplementary Materials). The agent in this model is a single decentralized individual. Each agent is characterized by phenotype and genotype. Main phenotypic parameters are: sex (male/female), hearing status (deaf/hearing) and sign language (knowledge/ignorance). The genotype is represented by one locus / gene in which a recessive allele is pathogenic. The main algorithm of the program represents life cycle of one generation (which considered equal to 20 years) of the model population. One cycle of the program includes: choice of marital partners based on phenotype; creation of a new population consisting of a progeny of agents of the current generation; modeling of consolidated communities of deaf people. We run the model in three different scenarios (combinations of initial parameters of the model population) in order to simulate the DFNB1A prevalence under different intensity of selection pressure (Supplementary Table 1). For each generation, the program registers data on the total population number, the number of deaf individuals, calculates the proportion of recessive mutant homozygotes and the frequency of recessive mutant allele, and compiles these parameters into the tables.

As a reference population for the developed model, we applied data on the Yakut population. The Yakuts (originally named as Sakha) are the largest indigenous people of Siberia (466,492 according to the Russian Census, 2010) living in the Sakha Republic (Eastern Siberia, Russia). The Yakuts are characterized by specific anthropological, demographic, linguistic and historical features indicated to their relationships to nomadic Turkic tribes of South Siberia and Central Asia. The genetic data revealed a relatively small size of Yakut ancestor population and the strong bottleneck effect in the Yakut paternal lineages (∼ 80% of Y chromosomes of Yakuts belong to one haplogroup - N3) [28]. Marriage traditions and geographical isolation had a significant role in genetic and demographic history of the Yakut population. High frequency of some Mendelian disorders in the Yakut population was found to be a result of the founder effect. For example, the high prevalence of HL in Yakuts is caused by the founder c.-23+1G>A mutation in the *GJB2* gene (92.2% of all mutant *GJB2* alleles found in deaf patients) and which was found with extremely high carrier frequency among hearing Yakut individuals (10.3% in total population) [17, 29]. Moreover, the data on marriage structure and reproduction of deaf people living in the Sakha Republic were presented in comparison with contribution of the *GJB2* gene mutations to the etiology of HL. The relative fertility of deaf people compared to their hearing siblings was 0.78 (mean number of children 1.76 and 2.24 to deaf and their hearing siblings, respectively). The rate of assortative marriages among deaf people was 77.1% [27]. The known genetic structure of hereditary HL in Yakuts and available data on reproductive capabilities and marital structure of deaf people make this population suitable for computer simulations of the distribution of DFNB1A.

The first scenario, “No deaf community”, was a model of population where deaf individuals did not mate and had no progeny, hence representing full pressure of “purifying” selection against deafness. The simulation results showed an increase in the population size from the initial 200,000 to 1,604,123 individuals in the 19^th^ generation, and the number of deaf individuals increased from the initial 530 to resulting 2,271 (Supplementary Table 2, Supplementary Figure 2). Accordingly, the frequency of recessive mutant allele decreased from 0.0542 to 0.0376 (Figure 1A, Supplementary Table 2) after 20 generations. The proportion of deaf individuals (recessive mutant homozygotes) slightly increased from 0.0027 to 0.0028 on the 1st generation, and then continuously decreased to 0.0014 by the 19^th^ generation (Figure 2B, Supplementary Table 2).

**Figure 2.**
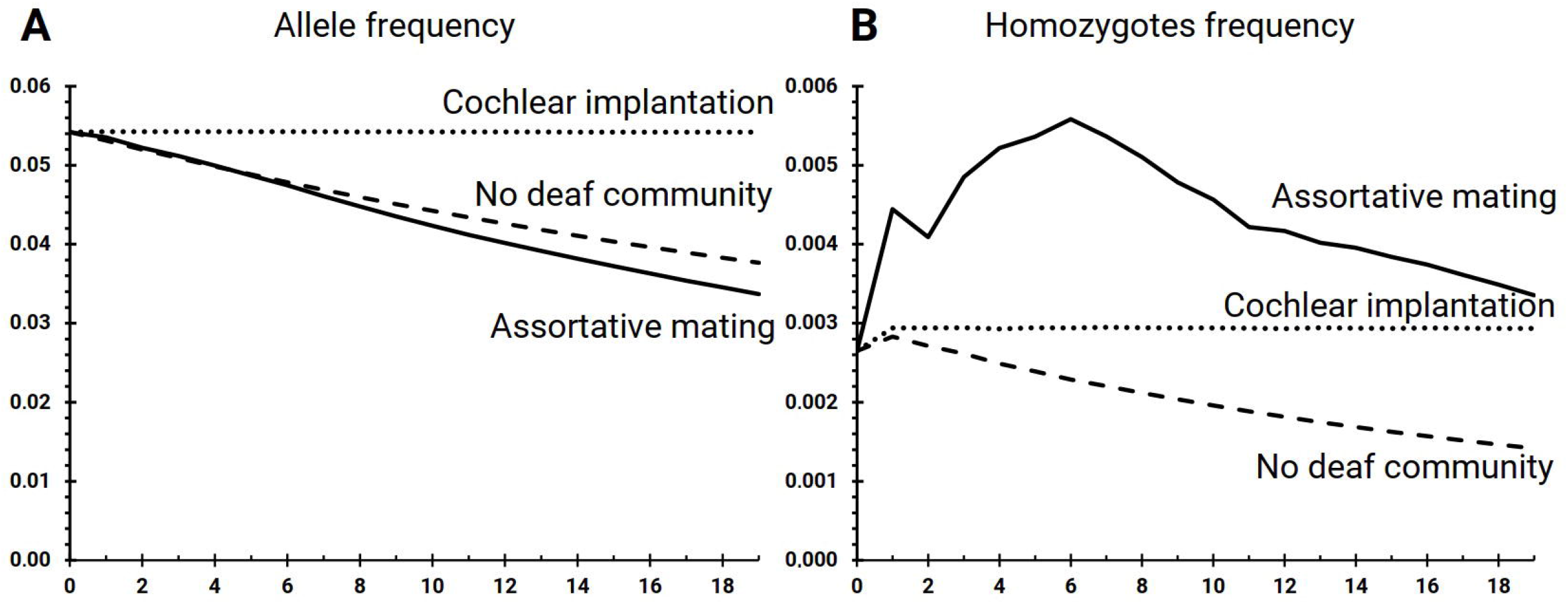
The simulation results for different scenarios. **A** – Frequency of recessive mutant allele; **B** – Proportion of deaf individuals (recessive mutant homozygotes). Y-axis: proportion; X-axis: generations (1 generation = 20 years).

The second scenario, “Assortative mating”, was a model of population where all deaf individuals used sign language for communication, and all marriages among them were assortative. This scenario represented relaxed selection due to the presence of linguistic homogamy among deaf individuals. The simulation results showed that the population number increased from initial 200,000 to 1,568,752 individuals in the 19^th^ generation, and the number of deaf individuals increased from initial 530 to resulting 5,320 (Supplementary Table 3, Supplementary Figure 3). Accordingly, the frequency of recessive mutant allele decreased from 0.0542 to 0.0337 after 20 generations (Figure 2A, Supplementary Table 3). The proportion of recessive mutant homozygotes increased up to the 6^th^ generation from 0.0027 to 0.0056, and then decreased to 0.0034 by the 19^th^ generation (Figure 1B, Supplementary Table 3).

The third scenario, “Cochlear implantation”, was a model of population where all deaf individuals acquire the ability to hear despite their pathogenic genotypes. This scenario represented neutral selection pressure due to the seeming lack of deafness phenotype. By this scenario, the population number increased from initial 200,000 to 1,653,731 individuals in the 19^th^ generation and the number of deaf individuals increased from initial 530 to resulting 4,853 (Supplementary Table 4, Supplementary Figure 4). The frequency of recessive mutant allele (0.0542) did not change, from the start to the 19^th^ generation (Figure 2A, Supplementary Table 4). The proportion of recessive mutant homozygotes slightly increased from 0.0027 to 0.0029 on the 1st generation, and then it was constant (0.0029) until the 19^th^ generation (Figure 2B, Supplementary Table 4). Total population number dynamics between all three scenarios were comparable. The number and proportion of deaf individuals (recessive mutant homozygotes) and the frequency of recessive allele changed differently in each scenario depending on intensity of modelled selection pressure.

## DISCUSSION

In this work, we have developed a computer model for analysis of the spread of hereditary congenital HL in an isolated human population. In order to test the different levels of selection pressure on deafness, we ran the program under three different scenarios (sets of initial parameter combinations of the model population). The developed models’ main algorithm is the mechanism for choosing a marital partner based on mutual assessment of agents depending on their phenotypic parameters – hearing or deaf, knowledge or ignorance of sign language.

The model population of the “No deaf community” scenario, showed a decrease of the proportion of deaf individuals (from 0.0027 to 0.0014) and the frequency of pathogenic allele (from 0.0542 to 0.0376) (Figure 2, Supplementary Table 2). This scenario assumed that deaf people cannot marry unless they use sign language to communicate (linguistic homogamy) and therefore cannot have offspring. Thus, the genetic fitness of deaf individuals is close to zero, which indicates a high selection pressure against deafness. According to this scenario, deaf children can be born (with a 25% probability) only from hearing parents who are both heterozygous carriers of a recessive pathogenic allele. Consequently, the observed continuous decrease of mutant allele frequency may be the result of a decreasing proportion of recessive mutant homozygotes in a population.

When assortative marriages are present, the proportion of deaf individuals (recessive mutant homozygotes) practically doubled (from 0.27% to 0.56%) during the first 6 generations (120 years), and then decreased to 0.34% in the 19^th^ generation (380 years), which is 2.42 times higher compared to the population without a deaf community (0.14%) (Figure 2, Supplementary Tables 2 and 3). The frequency of recessive mutant allele decreased from 0.0542 to 0.0337 (Figure 1A, Supplementary Table 3). These data suggest that the assortative marriages between deaf people based on linguistic homogamy (sign language) lead to an increase in the frequency of hereditary HL, as previously shown in other studies of the influence of social factors on hereditary HL (Figure 2) [24-26]. A decrease in the proportion of recessive mutant homozygotes after the 6th generation, shown in a population with the assortative marriages among deaf individuals, is associated with a reduced fertility of deaf individuals relative to hearing people by 22%. Such difference in fertility was set in the initial parameters of scenarios to represent actual data on the reproduction of deaf people in Yakutia [27]. Nevertheless, even with reduced reproductive capabilities, the proportion of the deaf individuals on the 19^th^ generation (after 380 years) is higher than in the absence of assortative marriages.

Of interest is a prolonged decrease of mutant allele frequency in a total population in both “No deaf community” and “Assortative mating” scenarios, from initial 0.0542 to 0.0376 and 0.0337, respectively, observed in this work (Figure 2A, Supplementary Tables 2 and 3). Regarding the “No deaf community” scenario, the frequency of mutant allele declined due to the absence of marriages between deaf individuals. Although, on the last generation, “assortative mating” scenario presented even lower frequency of mutant allele, which was 0.39% less than in “no deaf community” scenario. In a population in which HL is only due to recessive single-locus deafness (there are no other causes of HL), a couple of married deaf individuals will be able to have only deaf children, thereby making an increased contribution of the recessive mutant homozygotes to the next generation. A similar effect was observed in the study by Braun et al. (2020), where the proportion of homozygotes increased by 23% (from 0.017% to 0.022%) while the frequency of recessive pathogenic allele did not change (an increase of only 0.002%) [30].

The modeling according to the “Cochlear implantation” scenario showed that the prevalence of hereditary HL did not change, in contrast to the “Assortative mating” scenario (Figure 2, Supplementary Table 4, Supplementary Figure 4). This scenario presumed that all deaf individuals with cochlear implants “become” phenotypically hearing, despite their pathogenic genotypes. In this case there are no assortative marriages by deafness, since all individuals become “hearing” and genetic fitness of all individuals becomes equal regardless of their genotype. Therefore, the proportion of recessive homozygotes (*q*^2^) in the population will determine the probability of marriage of two deaf individuals (*q*^2^ × *q*^2^). And the proportion of such marriages will be much lower than in population with assortative mating by deafness. Thus, this scenario represents a panmictic population in which all individuals have equal genetic fitness, and the proportions of genotypes and allele frequencies will constant from generation to generation according to the Hardy-Weinberg principle.

Thus, the results of the modeling showed that in an isolated population with a low effective size and a high carrier frequency of pathogenic recessive allele the introduction of total cochlear implantation cannot lead to an increase in prevalence of hereditary HL (Figure 3). A similar “panmictic” scenario versus a scenario with assortative marriages by deafness were tested in two previous studies [24, 30]. Nance & Kearsey (2004) modelled a population with a totally random choice of a partner (random mating) and the equal reproductive capabilities of deaf and hearing individuals, which resulted in a minimal increase in the proportion of mutant homozygotes (by ∼1.5%) over 400 years [24]. Brown et al. (2020) showed that in a model population with random mating, the frequency of the pathogenic allele and the proportion of mutant homozygotes did not change over 200 years (10 generations) [30].

**Figure 3.**
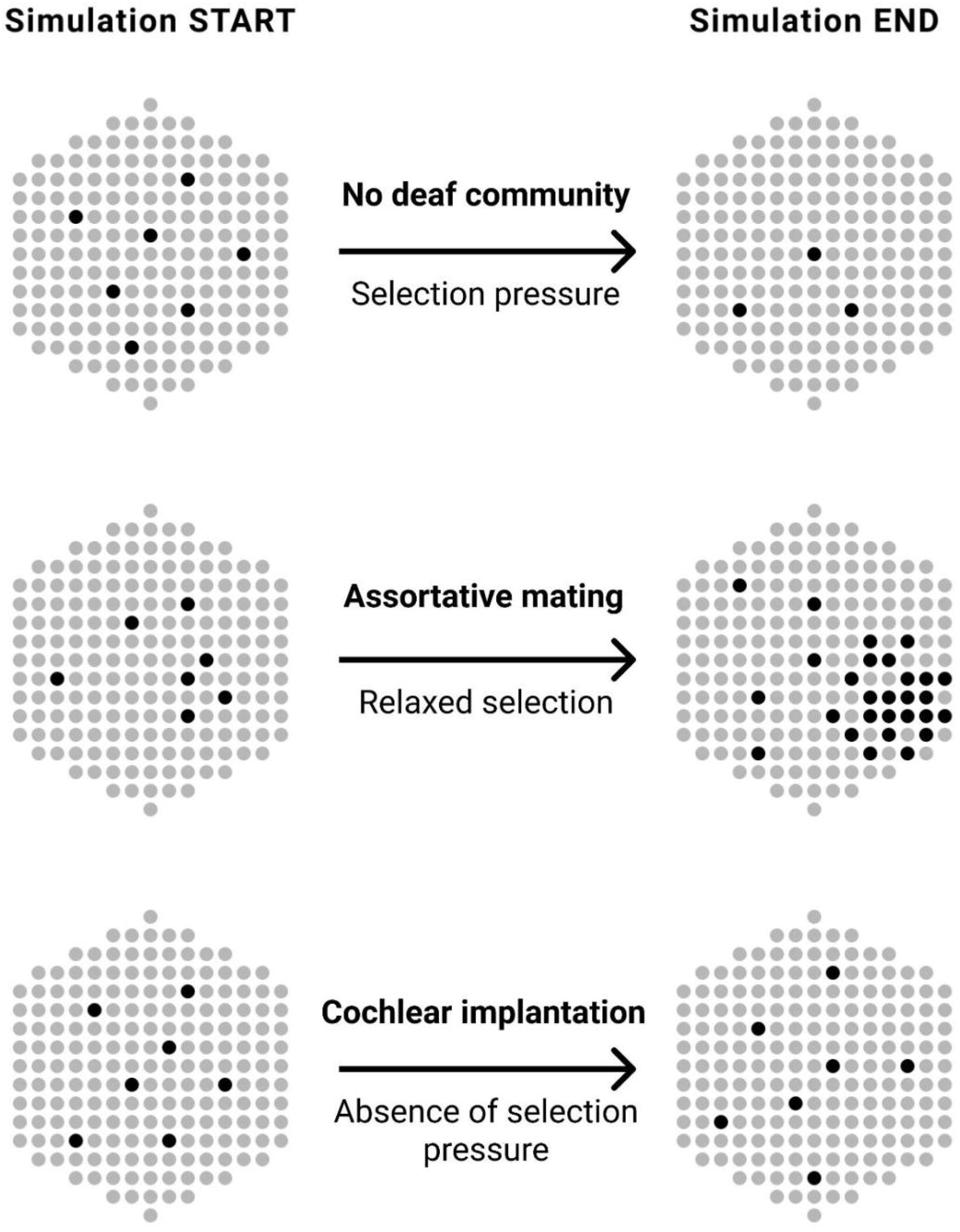
Visualization of the modeling results of three scenarios. The initial state of the model population is on the left; the final state (after 20 generations) is in the right. **A** – Scenario “No deaf community”. The decrease in the proportion of the deaf individuals was registered. **B** – Scenario “Assortative mating”. The increase in the proportion of the deaf individuals was registered. The use of sign language leads to assortative marriages among deaf individuals, forming the deaf community. **C** – Scenario “Cochlear implantation”. No differences in the HL incidence was registered due to the random mating pattern of all individuals.

The modeling results have shown that an initially low genetic fitness of deaf people can be greatly increased by assortative mating by deafness, resulting in a greater prevalence of DFNB1A. Contrary, in a model population where all deaf individuals undergo cochlear implantation, the initial frequencies of pathogenic allele and the prevalence of hereditary HL become constant.

## Data Availability

All data generated or analyzed during this study are included in this published article and its supplementary files

## DECLARATIONS

### Authors’ contributions

All authors read and approved the final manuscript. GPR – study design, project coordination, data interpretation and analysis, manuscript writing; AAS, VIZ – data interpretation and analysis, additional C++ and main python code writing; AMM, FVK – code review, manuscript proofreading; VGP, FMT, AVS – data interpretation and analysis, manuscript proofreading; SAF – project supervision, funding acquisition, manuscript proofreading; OLP – project supervision, data interpretation and analysis, manuscript proofreading; SAL – project supervision, study design, main C++ code writing and review, manuscript proofreading; NAB – project supervision, funding acquisition, study design, data interpretation and analysis, manuscript writing.

## Acknowledgements

The authors would like to express their sincere gratitude to Igor Dyachenko and Stepan Kutsenogiy for their participation in the project on its early stages.

## SUMMARY

It was evidenced, that the increase in the prevalence of autosomal recessive deafness 1A (DFNB1A) in populations of European descent was promoted by assortative marriages among deaf people. Sign language is the main type of communication of people with congenital hearing loss (HL). A widespread introduction of sign language has increased the genetic fitness of deaf individuals, thus relaxing selection against deafness. Whether relaxed selection will have a similar effect in populations with different genetic structures remains unclear. Currently, cochlear implantation is becoming a common method of rehabilitation for deaf patients, restoring their hearing ability and promoting the acquirement of spoken language. Whether the mass cochlear implantation could affect the spread of hereditary deafness is unknown. The aim of this study is a computer modeling of the DFNB1A prevalence in an isolated human population with regard to the genetic fitness of deaf individuals. We have developed an agent-based computer model for analysis of the spread of hereditary congenital recessive HL in an isolated human population. We run the model in three different combinations of initial parameters (scenarios) in order to model the DFNB1A prevalence under the different intensity of selection pressure for 20 generations (400 years). The “No deaf community” scenario represented the pressure of “purifying” selection on deafness and showed the decrease of the proportion of deaf individuals (from 0.0027 to 0.0014) and the pathogenic allele frequency (from 0.0542 to 0.0376). The “Assortative mating” scenario represented relaxed selection and showed the increase of the proportion of deaf individuals (from 0.0027 to 0.0034) and the decrease of the pathogenic allele frequency (from 0.0542 to 0.0337). The “Cochlear implantation” scenario representing neutral selection pressure, did not reveal significant changes in both the proportion of deaf individuals (0.0029) and the pathogenic allele frequency (0.0542). The modeling results have shown that the initially low genetic fitness of deaf people can be significantly increased in the presence of assortative mating by deafness, resulting in a higher prevalence of DFNB1A. Contrary, the modeling of a population where all deaf individuals undergo cochlear implantation showed that the initial frequency of pathogenic allele and the incidence of hereditary HL become constant.

## REFERENCES

1. Korver AMH, Smith RJH, Van Camp G, Schleiss MR, Bitner-Glindzicz MAK, Lustig LR, Usami S-i, Boudewyns AN: Congenital hearing loss. Nat Rev Dis Primers. 2017, 3(1). https://doi.org/10.1038/nrdp.2016.94.

2. Morton CC, Nance WE: Newborn Hearing Screening — A Silent Revolution. N Engl J Med. 2006, 354(20):2151–64. https://doi.org/10.1056/NEJMra050700.

3. Toriello H, Smith S: Hereditary Hearing Loss and Its Syndromes, 3 edn. Oxford: Oxford University Press; 2013.

4. Hereditary Hearing Loss Homepage https://hereditaryhearingloss.org. Accessed 30 Jul 2021.

5. Chan DK, Chang KW: GJB2-associated hearing loss: Systematic review of worldwide prevalence, genotype, and auditory phenotype. Laryngoscope. 2014, 124(2):E34–E53. https://doi.org/10.1002/lary.24332.

6. Stenson PD, Mort M, Ball EV, Evans K, Hayden M, Heywood S, Hussain M, Phillips AD, Cooper DN: The Human Gene Mutation Database: towards a comprehensive repository of inherited mutation data for medical research, genetic diagnosis and next-generation sequencing studies. Hum Genet. 2017, 136(6):665–677. https://doi.org/10.1007/s00439-017-1779-6.

7. Brobby GW, Müller-Myhsok B, Horstmann RD: Connexin 26 R143W Mutation Associated with Recessive Nonsyndromic Sensorineural Deafness in Africa. N Engl J Med. 1998, 338(8): 548–50. https://doi.org/10.1056/NEJM199802193380813.

8. Van Laer L: A common founder for the 35delG GJB2 gene mutation in connexin 26 hearing impairment. J Med Genet. 2001, 38(8):515–8. https://doi.org/10.1136/jmg.38.8.515

9. Shahin H, Walsh T, Sobe T, Lynch E, King M-C, Avraham KB, Kanaan M: Genetics of congenital deafness in the Palestinian population: multiple connexin 26 alleles with shared origins in the Middle East. Hum Genet. 2002, 110(3):284–9. https://doi.org/10.1007/s00439-001-0674-2.

10. RamShankar M: Contribution of connexin26 (GJB2) mutations and founder effect to non-syndromic hearing loss in India. J Med Genet. 2003, 40(5):e68. https://doi.org/10.1136/jmg.405.e68.

11. Yan D, Park H-J, Ouyang XM, Pandya A, Doi K, Erdenetungalag R, Du LL, Matsushiro N, Nance WE, Griffith AJ et al: Evidence of a founder effect for the 235delC mutation of GJB2 (connexin26) in east Asians. Hum Genet. 2003, 114(1):44–50. https://doi.org/10.1007/s00439-003-1018-1.

12. Tsukada K, Nishio S-y, Hattori M, Usami S-i: Ethnic-Specific Spectrum of GJB2 and SLC26A4 Mutations. Ann Otol Rhinol Laryngol. 2015, 124(1_suppl):61S–76S. https://doi.org/10.1177%2F0003489415575060.

13. Shinagawa J, Moteki H, Nishio S-y, Noguchi Y, Usami S-i: Haplotype Analysis of GJB2 Mutations: Founder Effect or Mutational Hot Spot? Genes. 2020, 11(3):250. https://doi.org/10.3390/genes11030250.

14. Wattanasirichaigoon D, Limwongse C, Jariengprasert C, Yenchitsomanus PT, Tocharoenthanaphol C, Thongnoppakhun W, Thawil C, Charoenpipop D, Phoiam T, Thongpradit S et al: High prevalence of V37I genetic variant in the connexin-26 (GJB2) gene among non-syndromic hearing-impaired and control Thai individuals. Clin Genet. 2004, 66(5):452–60. https://doi.org/10.1111/j.1399-0004.2004.00325.x.

15. Hamelmann C, Amedofu GK, Albrecht K, Muntau B, Gelhaus A, Brobby GW, Horstmann RD: Pattern of connexin 26 (GJB2) mutations causing sensorineural hearing impairment in Ghana. Hum Mutat. 2001, 18(1):84–5. https://doi.org/10.1002/humu.1156.

16. Morell RJ, Kim HJ, Hood LJ, Goforth L, Friderici K, Fisher R, Van Camp G, Berlin CI, Oddoux C, Ostrer H et al: Mutations in the connexin 26 gene (GJB2) among Ashkenazi Jews with nonsyndromic recessive deafness. N Engl J Med. 1998, 339(21):1500–5. https://doi.org/10.1056/NEJM199811193392103.

17. Barashkov NA, Dzhemileva LU, Fedorova SA, Teryutin FM, Posukh OL, Fedotova EE, Lobov SL, Khusnutdinova EK: Autosomal recessive deafness 1A (DFNB1A) in Yakut population isolate in Eastern Siberia: extensive accumulation of the splice site mutation IVS1+1G>A in GJB2 gene as a result of founder effect. J Hum Genet. 2011, 56(9):631–9. https://doi.org/10.1038/jhg.2011.72.

18. Carranza C, Menendez I, Herrera M, Castellanos P, Amado C, Maldonado F, Rosales L, Escobar N, Guerra M, Alvarez D et al: A Mayan founder mutation is a common cause of deafness in Guatemala. Clin Genet. 2016, 89(4):461–5. https://doi.org/10.1111/cge.12676.

19. Bliznetz EA, Lalayants MR, Markova TG, Balanovsky OP, Balanovska EV, Skhalyakho RA, Pocheshkhova EA, Nikitina NV, Voronin SV, Kudryashova EK et al: Update of the GJB2/DFNB1 mutation spectrum in Russia: a founder Ingush mutation del(GJB2-D13S175) is the most frequent among other large deletions. J Hum Genet. 2017, 62(8):789–95. https://doi.org/10.1038/jhg.2017.42.

20. Zytsar MV, Barashkov NA, Bady-Khoo MS, Shubina-Olejnik OA, Danilenko NG, Bondar AA, Morozov IV, Solovyev AV, Danilchenko VY, Maximov VN et al: Updated carrier rates for c.35delG (GJB2) associated with hearing loss in Russia and common c.35delG haplotypes in Siberia. BMC Med Genet. 2018, 19(1). https://doi.org/10.1186/s12881-018-0650-5.

21. Posukh OL, Zytsar MV, Bady-Khoo MS, Danilchenko VY, Maslova EA, Barashkov NA, Bondar AA, Morozov IV, Maximov VN, Voevoda MI: Unique Mutational Spectrum of the GJB2 Gene and Its Pathogenic Contribution to Deafness in Tuvinians (Southern Siberia, Russia): A High Prevalence of Rare Variant c.516G>C (p.Trp172Cys). Genes. 2019, 10(6):429. https://doi.org/10.3390/genes10060429.

22. Zytsar MV, Bady-Khoo MS, Danilchenko VY, Maslova EA, Barashkov NA, Morozov IV, Bondar AA, Posukh OL: High Rates of Three Common GJB2 Mutations c.516G>C, c.-23+1G>A, c.235delC in Deaf Patients from Southern Siberia Are Due to the Founder Effect. Genes. 2020, 11(7):833. https://doi.org/10.3390/genes11070833.

23. Nance WE, Liu X-Z, Pandya A: Relation between choice of partner and high frequency of connexin-26 deafness. Lancet. 2000, 356(9228):500–1. https://doi.org/10.1016/S0140-6736(00)02565-4.

24. Nance WE, Kearsey MJ: Relevance of Connexin Deafness (DFNB1) to Human Evolution. Am J Hum Genet. 2004, 74(6):1081–7. https://doi.org/10.1086/420979.

25. Arnos KS, Welch KO, Tekin M, Norris VW, Blanton SH, Pandya A, Nance WE: A comparative analysis of the genetic epidemiology of deafness in the United States in two sets of pedigrees collected more than a century apart. Am J Hum Genet. 2008, 83(2):200–7. https://doi.org/10.1016/j.ajhg.2008.07.001.

26. Blanton SH, Nance WE, Norris VW, Welch KO, Burt A, Pandya A, Arnos KS: Fitness Among Individuals with Early Childhood Deafness: Studies in Alumni Families from Gallaudet University. Ann Hum Genet. 2010, 74(1):27–33. https://doi.org/10.1111/j.1469-1809.2009.00553.x.

27. Romanov GP, Barashkov NA, Teryutin FM, Lashin SA, Solovyev AV, Pshennikova VG, Bondar AA, Morozov IV, Sazonov NN, Tomsky MI et al: Marital Structure, Genetic Fitness, and the GJB2 Gene Mutations among Deaf People in Yakutia (Eastern Siberia, Russia). Russ J Genet. 2018, 54(5):554–61. https://doi.org/10.1134/S1022795418050071.

28. Fedorova SA, Reidla M, Metspalu E, Metspalu M, Rootsi S, Tambets K, Trofimova N, Zhadanov SI, Hooshiar Kashani B, Olivieri A et al: Autosomal and uniparental portraits of the native populations of Sakha (Yakutia): implications for the peopling of Northeast Eurasia. BMC Evol Biol. 2013, 127(13). https://doi.org/10.1186/1471-2148-13-127.

29. Barashkov NA, Pshennikova VG, Posukh OL, Teryutin FM, Solovyev AV, Klarov LA, Romanov GP, Gotovtsev NN, Kozhevnikov AA, Kirillina EV et al: Spectrum and Frequency of the GJB2 Gene Pathogenic Variants in a Large Cohort of Patients with Hearing Impairment Living in a Subarctic Region of Russia (the Sakha Republic). PLoS One. 2016, 11(5):e0156300. https://doi.org/10.1371/journal.pone.0156300.

30. Braun DC, Jain S, Epstein E, Greenwald BH, Herold B, Gray M: Deaf intermarriage has limited effect on the prevalence of recessive deafness and no effect on underlying allelic frequency. PLoS One 2020, 15(11):e0241609. https://doi.org/10.1371/journal.pone.0241609

