## Supplementary materials for "Agent-based modeling of DFNB1A prevalence with regard to intensity of selection pressure in isolated human population: will cochlear implantation increase the cases of hereditary deafness?"

### Program algorithm

At the first iteration, the program generates the initial population which consists of the arrays "men" and "women" filled by the agents according to the specified numbers of men and women (parameters `INIT_POP_SIZE_M` and `INIT_POP_SIZE_W`). When each agent is generated, his/her genotype is specified using the parameters `alleleFather` and `alleleMother` which can take on the values 0 or 1, where 1 is the recessive mutant allele (deafness allele), and 0 is the dominant normal allele. The first allele (`alleleFather`), in turn, is specified as follows: if a random number from the uniform distribution from 0 to 1 is less than the value of `DEAF_ALLELE_RATIO` that we set, then the value of the `alleleFather` parameter will be equal to 1. The value of the second parameter (`alleleMother`) will be equal to 1 if the condition `alleleFather` = 1 is true and a random number from the uniform distribution from 0 to 1 is less than or equal to the value of `DEAF_HOMOZYGOTES` that we set. Otherwise `alleleMother` will be equal to 0.

To determine whether new agent will have deafness phenotype, the sum of the values of the `alleleFather` and `alleleMother` parameters is determined and if it is greater than 1, then the agent is considered to be deaf (the value of the `isDeaf` parameter is set to `true`), otherwise the agent is considered to be hearing (the value of the `isDeaf` parameter is set to `false`).

Then the program specifies whether the agent will have acquired (non-hereditary) deafness similar to the setting of the previous parameters - if a random number from a uniform distribution is less than the value of the `SPONTANEOUS_DEAF` parameter that we specified, then the agent will be deaf, regardless of genotype.

Next, we specify the value of the agent's `socialPosition` parameter which is the minimum required sum of the values of the estimated parameters of the partner for the possibility of marriage. The value of the `socialPosition` parameter is set as a random number from the normal distribution with the mean equal to the `SOCIAL_MEAN_D` parameter and the standard deviation equal to `SOCIAL_VAR_D` if the agent is deaf. If the agent is hearing, then the mean and standard deviation are equal to `SOCIAL_MEAN_H` and `SOCIAL_VAR_H`, respectively.

Whether the agent will know the sign language is specified by a random value from a uniform distribution if it is less than `0.5`.

For the 2nd and all subsequent iterations:

Starting from the 2nd iteration, the new population consists exclusively of descendants from the previous iteration. All parameters of the agent described above are specified in a different way, described in the section "Algorithm for generating offspring".

After the creation of the population, the main process starts - the formation of couples and the birth of children.

**Algorithm for mutual assessment of partners for marriage**

To simulate the process of marriage of agents, a matrix is formed, the rows of which correspond to male agents and the columns to female agents (Supplementary Figure 1). Each cell of this matrix contains the so-called "score" `s` assigned to each potential couple according to the algorithm for mutual evaluation of partners. Next, line by line, for each male agent the program selects a female agent with the highest "score"; a couple is created and the offspring is generated, after which the selected female agent becomes inaccessible to other candidates.

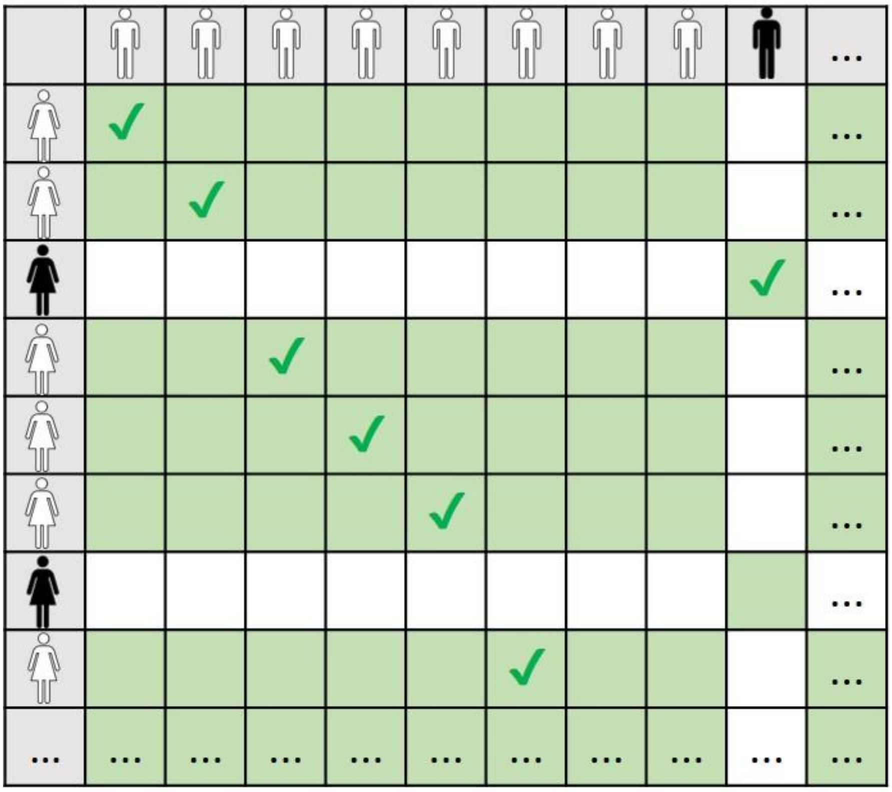

**Supplementary Figure 1. The scheme of mutual assessment of candidate partners in case of the "assortative mating" scenario.** Check marks – marriage. Green cells – marriage considered. White cells – marriage not considered.

The first step is to check if the candidates are brother and sister. For this, the values of the `fatherID` and `motherID` parameters of each candidate are compared, and if any of them coincide, the mutual evaluation is not conducted and the creation of such a couple becomes impossible.

If the candidates are not brothers and sisters, then the calculation of the `S` score begins according to the following rules:

1. If candidates have the same phenotype (both hearing or both deaf), then:

1.1. If both candidates are hearing, the values of the WEIGHT_PHENO_H parameters of both candidates are added to the S score (i.e. 2 × WEIGHT_PHENO_H).

1.2. If both candidates are deaf, the value 2 × WEIGHT_PHENO_H is added to the S score. Also added to the S score is the value WEIGHT_SIGN_D, if one of the candidates knows sign language, or 2 × WEIGHT_SIGN_D, if both candidates know sign language.

2. If candidates differ in phenotype (one is deaf, the other is hearing), then the sum (WEIGHT_SIGN_H + WEIGHT_SIGN_D) will be added to the S score only if both candidates know sign language.

After the S score is calculated, the program compares it with the sum of the socialPosition values obtained for each candidate. And if the S score exceeds the sum of the socialPosition thresholds, then the value of S is written in the cell of the matrix corresponding to the given pair of candidates.

**Algorithm for generating offspring**

If the marriage between the candidates took place, then the process of generating offspring begins.

First, the type of couple is determined based on the phenotypes of the partners (DD, DH or HH, where D is deaf, H is hearing). For couples of HH type, the value of the BIRTH_RATE_H parameter is used in further calculations; for couples of DD and DH types, the value of the BIRTH_RATE_D parameter is used. Next, the number of children for the current couple is determined using the beta distribution with the BETA_A and BETA_B parameters, the values of which are adjusted according to the BIRTH_RATE_D or BIRTH_RATE_H parameters, depending on the type of marriage. Based on the generated number of descendants, the program creates the corresponding number of new agents which are equally likely to be male or female.

The genotype of a new agent is formed based on the genotypes of the father and mother and is stored in the parameters mGenome (allele inherited from the mother) and fGenome (allele inherited from the father). The agent will be deaf for genetic reasons only if both alleles in the genotype are recessive, in which case the isDeaf parameter will have the value true. Whether the deafness of an agent is acquired is determined in the same way, as described above for the formation of the initial population. The knowsSignLanguage parameter (knowledge of sign language) is determined based on the family situation - if at least one deaf parent in the family knows sign language, then all children will automatically know it. The socialPosition parameter is determined as the arithmetic mean of the socialPosition parameters of the parents adjusted for some normally distributed value.

The following parameters are set in the model:

| | Parameter | Range of values | Description |
|---|---|---|---|
| 1 | MAX_GENERATIONS | 1≤x≤∞, x∈N | Number of generations |
| 2 | INIT_POP_SIZE_M | 1≤x≤∞, x∈N | Initial size of male population |
| 3 | INIT_POP_SIZE_W | 1≤x≤∞, x∈N | Initial size of female population |
| 4 | SPONTANEOUS_DEAF | 0≤x≤1, x∈R | Proportion of agents with spontaneous deafness due to non-genetic reasons |
| 5 | DEAF_ALLELE_RATIO | 0≤x≤1, x∈R | Proportion of mutant allele in initial population |
| 6 | DEAF_HOMOZYGOTES | 0≤x≤1, x∈R | Proportion of the deaf in initial population |
| 7 | BIRTH_RATE_H | 0≤x≤∞, x∈$N_0$ | Average number of children of hearing agent |
| 8 | BIRTH_RATE_D | 0≤x≤∞, x∈ $N_0$ | Average number of children of deaf agent |
| 9 | BETA_A, BETA_B | x>0 | Boundaries of beta distribution interval |
| 10 | SOCIAL_MEAN_H | 0≤x≤1, x∈R | Lower threshold of sum of parameter values of potential partner for *hearing* agent |
| 11 | SOCIAL_VAR_H | 0≤x≤1, x∈R | Standard deviation of normal distribution for calculating SOCIAL_MEAN_H of specific *hearing* agent |
| 12 | SOCIAL_MEAN_D | 0≤x≤1, x∈R | Lower threshold of sum of parameter values of potential partner for *deaf* agent |
| 13 | SOCIAL_VAR_D | 0≤x≤1, x∈R | Standard deviation of normal distribution for calculating SOCIAL_MEAN_D of specific *deaf* agent |
| 14 | WEIGHT_PHENO_H | 0≤x≤1, x∈R | Significance of phenotype match with potential partner for *hearing* agent |
| 15 | WEIGHT_SIGN_H | 0≤x≤1, x∈R | Significance of sign language knowledge by potential partner for *hearing* agent |
| 16 | WEIGHT_PHENO_D | 0≤x≤1, x∈R | Significance of phenotype match with potential partner for *deaf* agent |
| 17 | WEIGHT_SIGN_D | 0≤x≤1, x∈R | Significance of sign language knowledge by potential partner for *deaf* agent |

When generated, each agent receives the following set of parameters:

| | Parameter | Range of values | Description |
|---|---|---|---|
| 1 | mGenome | {0, 1}<br>(1 – recessive allele, 0 - dominant) | Allele inherited from mother |
| 2 | fGenome | {0, 1}<br>(1 – recessive allele, 0 - dominant) | Allele inherited from father |
| 3 | isDeaf | true/false | Deafness of agent (true=deaf) |
| 4 | knowsSignLanguage | true/false | Knowledge of sign language |
| 5 | socialPosition | 0≤x≤1, x∈R | Lower threshold of sum of parameter |

| | | | |
|---|---|---|---|
| | | | values of potential partner |
| 6 | fatherID | ? | Agent father ID |
| 7 | motherID | ? | Agent mother ID |

As a result, the program generates a table with the values of the parameters of interest by generations.

**Supplementary Table 1. Initial simulation parameters for different scenarios.**

| No | Parameter | Scenario 1 “No deaf community” | Scenario 2 “Assortative mating” | Scenario 3 “Cochlear implantation” | Verification scenario |
|---|---|---|---|---|---|
| 1 | generations | 20; | 20; | 20; | 20; |
| 2 | init_pop_men | 100000; | 100000; | 100000; | 100000; |
| 3 | init_pop_women | 100000; | 100000; | 100000; | 100000; |
| 4 | candidate_pairs_mean | 50; | 50; | 50; | 50; |
| 5 | candidate_pairs_var | 15; | 15; | 15; | 15; |
| 6 | beta_ad | 100.0f; | 100.0f; | 100.0f; | 100.0f; |
| 7 | beta_bd | 100.0f; | 100.0f; | 100.0f; | 100.0f; |
| 8 | spontaneous_deaf | 0.00f; | 0.00f; | 0.00f; | 0.00f; |
| 9 | deaf_homozygotes | 0.002652f; | 0.002652f; | 0.002652f; | 0.0002f; |
| 10 | hear_homozygotes | 0.8943f; | 0.8943f; | 0.8943f; | 0.9715f; |
| 11 | birth_rate_hearing | 2.24f; | 2.24f; | 2.24f; | 2.24f; |
| 12 | birth_rate_deaf | 1.76f; | 1.76f; | 2.24f; | 2.016f; |
| 13 | birth_rate_mixed | 2.0f; | 2.0f; | 2.24f; | 2.128f; |
| 14 | beta_a | 2.0f; | 2.0f; | 2.0f; | 2.0f; |
| 15 | beta_b | 5.0f; | 5.0f; | 5.0f; | 5.0f; |
| 16 | social_mean_h | 0.55f; | 0.55f; | 0.3f; | 0.55f; |
| 17 | social_mean_d | 0.55f; | 0.55f; | 0.3f; | 0.55f; |
| 18 | social_var_h | 0.0f; | 0.0f; | 0.0f; | 0.0f; |
| 19 | social_var_d | 0.0f; | 0.0f; | 0.0f; | 0.0f; |
| 20 | weight_phenotype_hearing | 0.6f; | 0.6f; | 0.4f; | 0.6f; |
| 21 | weight_phenotype_deaf | 0.3f; | 0.3f; | 0.0f; | 0.3f; |
| 22 | weight_sign_h | 0.6f; | 0.6f; | 0.4f; | 0.6f; |
| 23 | weight_sign_d | 0.3f; | 0.3f; | 0.4f; | 0.3f; |
| 24 | sign_lang_d | 0.0f; | 1.0f; | 1.0f; | 1.0f; |
| 25 | sign_lang_h | 0.0f; | 0.0f; | 1.0f; | 0.0f; |
| 26 | deaf_community_model | 0; | 1; | 0; | 1; |

**Supplementary Table 2. Simulation results for scenario 1 "No deaf community".**

| Years | Total Population (0.99 CI) | Deaf individuals (0.99 CI) | Allele frequency (0.99 CI) | Genotype frequency (0.99 CI) |
|---|---|---|---|---|
| 0 | 200000.00 (±0) | 530.00 (±0) | 0.0542 (±0) | 0.0027 (±0) |
| 20 | 223198.33 (±0.25) | 632.01 (±0.59) | 0.0531 (±0.00) | 0.0028 (±0.00) |
| 40 | 248957.40 (±8.31) | 675.48 (±1.08) | 0.0520 (±0.00) | 0.0027 (±0.00) |
| 60 | 278034.45 (±1.83) | 727.46 (±1.27) | 0.0509 (±0.00) | 0.0026 (±0.00) |
| 80 | 309692.65 (±15.18) | 770.63 (±2.76) | 0.0498 (±0.00) | 0.0025 (±0.00) |
| 100 | 345209.04 (±7.31) | 825.22 (±0.31) | 0.0488 (±0.00) | 0.0024 (±0.00) |
| 120 | 384057.96 (±67.34) | 878.66 (±3.04) | 0.0478 (±0.00) | 0.0023 (±0.00) |
| 140 | 428242.58 (±27.70) | 943.68 (±1.52) | 0.0469 (±0.00) | 0.0022 (±0.00) |
| 160 | 477212.75 (±21.72) | 1011.19 (±2.73) | 0.0460 (±0.00) | 0.0021 (±0.00) |
| 180 | 533214.89 (±11.00) | 1086.00 (±1.27) | 0.0451 (±0.00) | 0.0020 (±0.00) |
| 200 | 595334.55 (±26.90) | 1165.90 (±2.36) | 0.0442 (±0.00) | 0.0020 (±0.00) |
| 220 | 664242.34 (±208.03) | 1252.77 (±1.18) | 0.0434 (±0.00) | 0.0019 (±0.00) |
| 240 | 741019.88 (±3.07) | 1345.57 (±1.55) | 0.0426 (±0.00) | 0.0018 (±0.00) |
| 260 | 828388.14 (±7.56) | 1447.47 (±1.36) | 0.0418 (±0.00) | 0.0017 (±0.00) |
| 280 | 923574.64 (±164.09) | 1557.57 (±2.54) | 0.0411 (±0.00) | 0.0017 (±0.00) |
| 300 | 1031186.17 (±62.66) | 1677.16 (±7.59) | 0.0403 (±0.00) | 0.0016 (±0.00) |
| 320 | 1150785.46 (±348.70) | 1809.05 (±3.35) | 0.0396 (±0.00) | 0.0016 (±0.00) |
| 340 | 1284825.00 (±63.41) | 1947.38 (±2.67) | 0.0389 (±0.00) | 0.0015 (±0.00) |
| 360 | 1435520.40 (±47.94) | 2101.42 (±1.92) | 0.0383 (±0.00) | 0.0015 (±0.00) |
| 380 | 1604123.34 (±361.31) | 2271.73 (±2.63) | 0.0376 (±0.00) | 0.0014 (±0.00) |

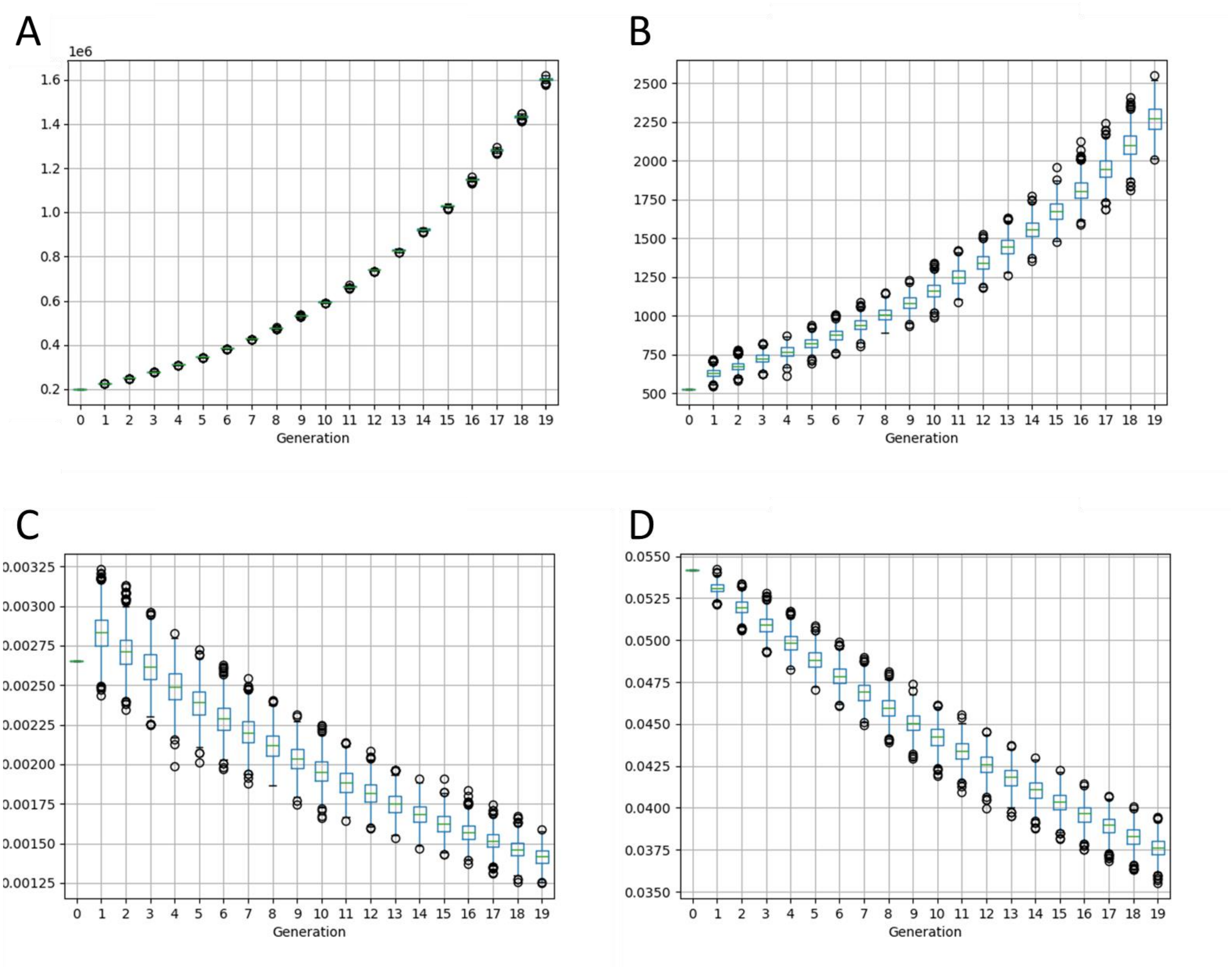


**Supplementary Figure 2**. Simulation results for scenario 1 – "No deaf community". **A** – Total population size. **B** – The number of deaf individuals. **C** – Proportion of recessive mutant homozygotes in total population. **D** – Frequency of recessive mutant allele in total population.

**Supplementary Table 3. Simulation results for scenario 2 “Assortative mating”.**

| Years | Total Population (0.99 CI) | Deaf individuals (0.99 CI) | Allele frequency (0.99 CI) | Genotype frequency (0.99 CI) |
|---|---|---|---|---|
| 0 | 200000.00 (±0) | 530.00 (±0) | 0.0542 (±0) | 0.0027 (±0) |
| 20 | 223294.15 (±0.56) | 992.19 (±0.06) | 0.0535 (±0.00) | 0.0044 (±0.00) |
| 40 | 249028.33 (±3.81) | 1018.78 (±2.01) | 0.0522 (±0.00) | 0.0041 (±0.00) |
| 60 | 278115.49 (±18.50) | 1348.71 (±1.83) | 0.0512 (±0.00) | 0.0048 (±0.00) |
| 80 | 309672.35 (±13.82) | 1615.56 (±2.73) | 0.0500 (±0.00) | 0.0052 (±0.00) |
| 100 | 344986.34 (±16.61) | 1849.88 (±2.91) | 0.0487 (±0.00) | 0.0054 (±0.00) |
| 120 | 383491.73 (±11.47) | 2141.06 (±3.16) | 0.0474 (±0.00) | 0.0056 (±0.00) |
| 140 | 427237.04 (±24.39) | 2291.74 (±4.43) | 0.0461 (±0.00) | 0.0054 (±0.00) |
| 160 | 475670.05 (±66.91) | 2427.23 (±10.13) | 0.0448 (±0.00) | 0.0051 (±0.00) |
| 180 | 531083.31 (±43.39) | 2540.98 (±2.51) | 0.0435 (±0.00) | 0.0048 (±0.00) |
| 200 | 592422.85 (±106.36) | 2704.67 (±9.33) | 0.0423 (±0.00) | 0.0046 (±0.00) |
| 220 | 660573.29 (±10.54) | 2784.80 (±4.12) | 0.0412 (±0.00) | 0.0042 (±0.00) |
| 240 | 736496.17 (±101.80) | 3069.83 (±2.23) | 0.0402 (±0.00) | 0.0042 (±0.00) |
| 260 | 822638.72 (±43.88) | 3305.75 (±2.05) | 0.0391 (±0.00) | 0.0040 (±0.00) |
| 280 | 916571.79 (±45.12) | 3625.28 (±4.71) | 0.0382 (±0.00) | 0.0040 (±0.00) |
| 300 | 1022709.73 (±56.84) | 3928.44 (±2.29) | 0.0372 (±0.00) | 0.0038 (±0.00) |
| 320 | 1140513.60 (±174.53) | 4268.56 (±14.66) | 0.0363 (±0.00) | 0.0037 (±0.00) |
| 340 | 1272681.96 (±8.89) | 4597.76 (±13.98) | 0.0354 (±0.00) | 0.0036 (±0.00) |
| 360 | 1420875.25 (±234.00) | 4956.84 (±6.17) | 0.0345 (±0.00) | 0.0035 (±0.00) |
| 380 | 1586752.25 (±209.86) | 5320.00 (±2.23) | 0.0337 (±0.00) | 0.0034 (±0.00) |

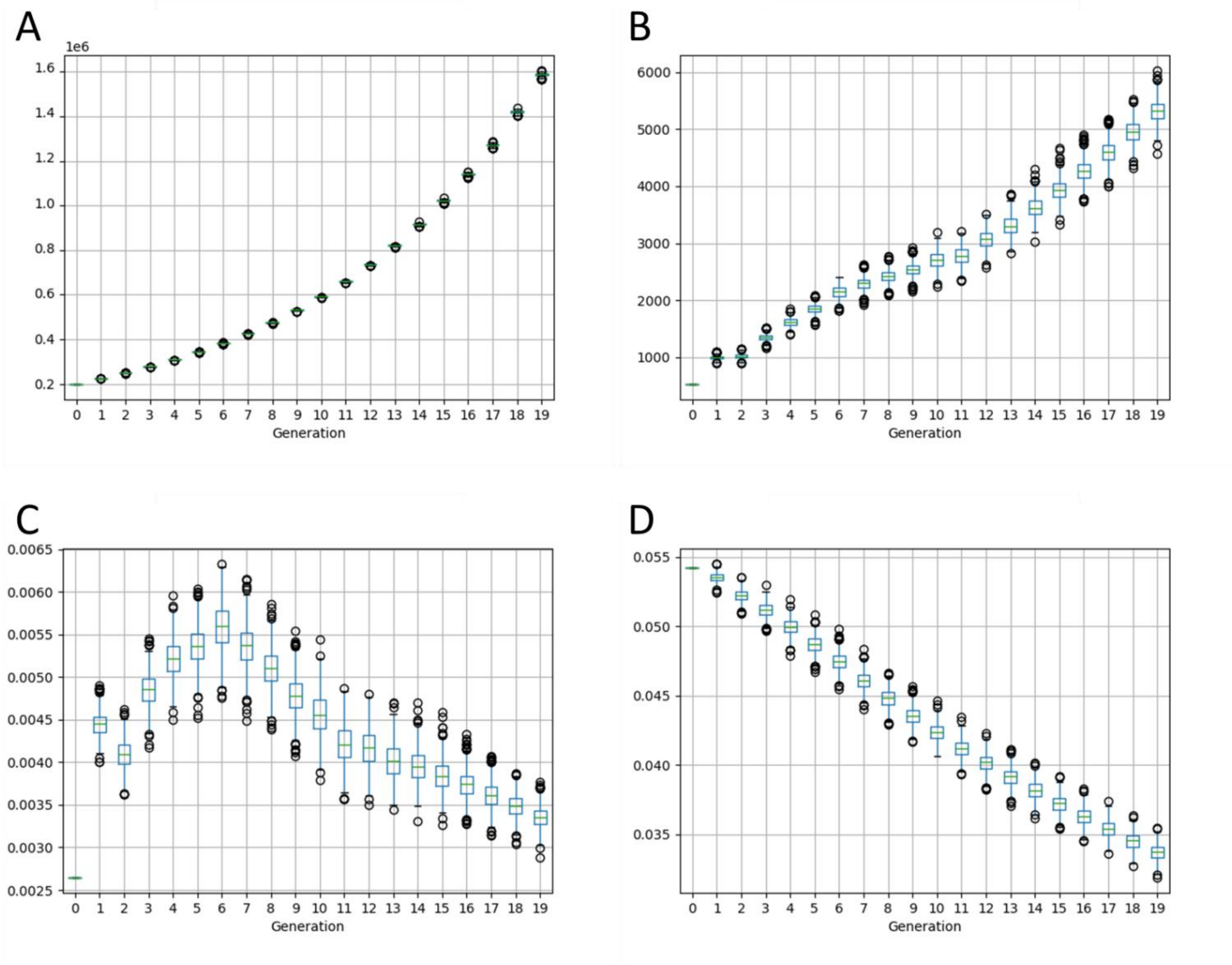


**Supplementary Figure 3.** Simulation results for scenario 2 – "Assortative mating". **A** – Total population size. **B** – The number of deaf individuals. **C** – Proportion of recessive mutant homozygotes in total population. **D** – Frequency of recessive mutant allele in total population.

**Supplementary Table 4. Simulation results for scenario 3 "Cochlear implantation".**

| Years | Total Population (0.99 CI) | Deaf individuals (0.99 CI) | Allele frequency (0.99 CI) | Genotype frequency (0.99 CI) |
|---|---|---|---|---|
| 0 | 200000.00 (±0) | 530.00 (±0) | 0.0542 (±0) | 0.0027 (±0) |
| 20 | 223625.69 (±1.12) | 657.45 (±0.65) | 0.0542 (±0.00) | 0.0029 (±0.00) |
| 40 | 249958.38 (±28.32) | 735.10 (±1.36) | 0.0542 (±0.00) | 0.0029 (±0.00) |
| 60 | 279783.58 (±7.41) | 823.03 (±2.57) | 0.0542 (±0.00) | 0.0029 (±0.00) |
| 80 | 312247.31 (±44.66) | 914.75 (±1.30) | 0.0542 (±0.00) | 0.0029 (±0.00) |
| 100 | 348761.81 (±1.30) | 1026.71 (±4.40) | 0.0542 (±0.00) | 0.0029 (±0.00) |
| 120 | 388568.18 (±66.44) | 1142.17 (±1.80) | 0.0542 (±0.00) | 0.0029 (±0.00) |
| 140 | 434236.79 (±63.47) | 1280.10 (±6.72) | 0.0542 (±0.00) | 0.0029 (±0.00) |
| 160 | 484570.86 (±24.45) | 1425.54 (±1.08) | 0.0542 (±0.00) | 0.0029 (±0.00) |
| 180 | 542343.30 (±126.66) | 1594.92 (±0.87) | 0.0542 (±0.00) | 0.0029 (±0.00) |
| 200 | 606807.31 (±116.12) | 1783.52 (±1.49) | 0.0542 (±0.00) | 0.0029 (±0.00) |
| 220 | 677732.35 (±7.56) | 1991.17 (±1.77) | 0.0542 (±0.00) | 0.0029 (±0.00) |
| 240 | 757188.70 (±44.94) | 2219.81 (±1.52) | 0.0542 (±0.00) | 0.0029 (±0.00) |
| 260 | 847587.96 (±63.93) | 2495.19 (±8.46) | 0.0542 (±0.00) | 0.0029 (±0.00) |
| 280 | 945984.77 (±110.88) | 2778.45 (±14.07) | 0.0542 (±0.00) | 0.0029 (±0.00) |
| 300 | 1057372.92 (±261.15) | 3101.98 (±8.96) | 0.0542 (±0.00) | 0.0029 (±0.00) |
| 320 | 1181858.34 (±125.60) | 3473.00 (±3.94) | 0.0542 (±0.00) | 0.0029 (±0.00) |
| 340 | 1321138.51 (±93.93) | 3880.55 (±7.31) | 0.0542 (±0.00) | 0.0029 (±0.00) |
| 360 | 1478374.62 (±253.65) | 4338.92 (±3.10) | 0.0542 (±0.00) | 0.0029 (±0.00) |
| 380 | 1653731.97 (±548.55) | 4853.76 (±3.25) | 0.0542 (±0.00) | 0.0029 (±0.00) |

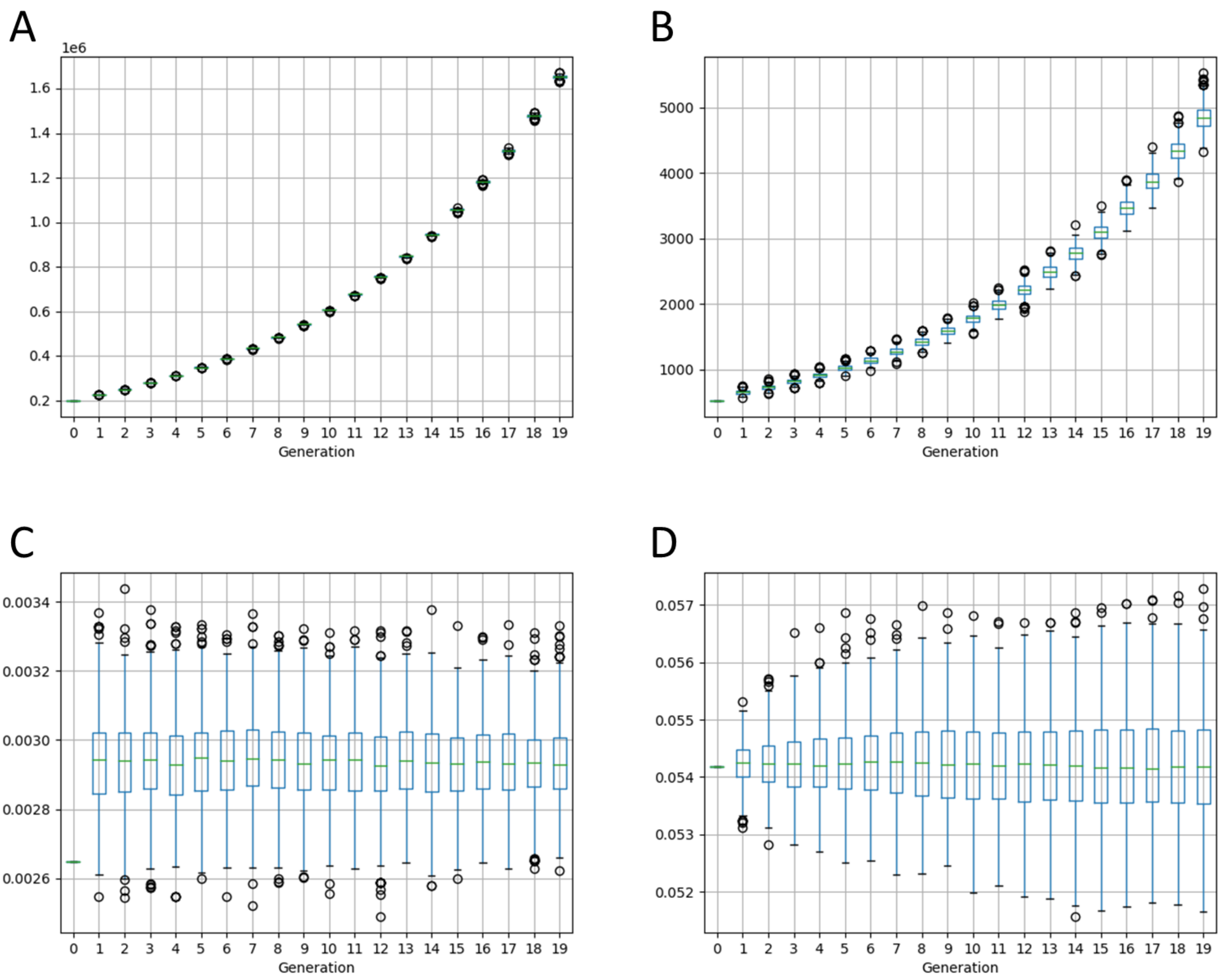


**Supplementary Figure 4.** Simulation results for scenario 3 “Cochlear implantation”. **A** – Total population size. **B** – The number of deaf individuals. **C** – Proportion of recessive mutant homozygotes in total population. **D** – Frequency of recessive mutant allele in total population.

#### Combinations of phenotypes and genotypes of marital partners

Possible combinations of marital partners according to their phenotypes are presented in the scheme (Supplementary Figure 5). In the absence of sign language, deaf individuals practically do not marry and, as can be seen from the scheme, a child with hereditary HL can only be born to a couple of hearing individuals who are heterozygous carriers of a recessive mutant allele. With the emergence of sign language, the marriages among the deaf become possible; the likelihood for them to have deaf children varies in different combinations of married couples with genetic and non-genetic causes of deafness.

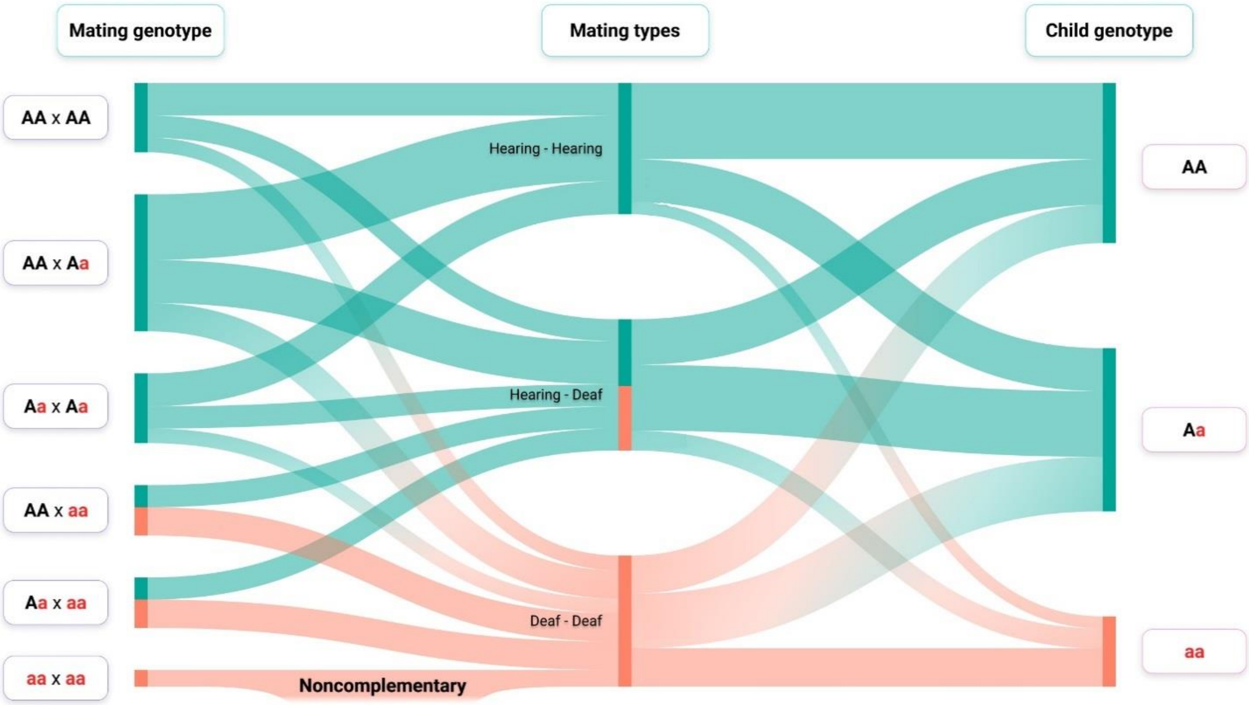


**Supplementary Figure 5. Combinations of marital partners by phenotype and genotype and possible genotypes in children.** On the left are genotypes of marital partners. In the middle are the marriages types by phenotypes of partners. On the right are possible genotypes in children of different marriages types. A – normal allele, a – mutant recessive allele.

An analysis of assortative marriages of deaf individuals, taking into account the ability to hear of their descendants, makes it possible to divide them into two groups - the so-called complementary and non-complementary assortative marriages [1]. Complementary marriages mean the marriages between deaf marital partners with different etiology of HL (acquired HL in one of the partners or mutations in different genes associated with HL); in such marriages, there may be only hearing or, in some cases, both deaf and hearing children. Non-complementary marriages are the marriages between deaf individuals who have the same genetic cause of HL – the presence of biallelic recessive mutations of the same gene. All children of such married couple will also be deaf and have the same genetic etiology of HL as their parents. From this category of marriages, the transmission of mutant alleles to the next generation is expected to be much higher than on average [1, 2]. The introduction of cochlear implantation leads to the emergence of new combinations of marital partners, both by phenotype and genotype (Supplementary Figure 5).

REFERENCES

1. Nance WE, Kearsey MJ: Relevance of Connexin Deafness (DFNB1) to Human Evolution. Am J Hum Genet. 2004, 74(6):1081-7. https://doi.org/10.1086/420979.
2. Arnos KS, Welch KO, Tekin M, Norris VW, Blanton SH, Pandya A, Nance WE: A comparative analysis of the genetic epidemiology of deafness in the United States in two sets of pedigrees collected more than a century apart. Am J Hum Genet. 2008, 83(2):200-7. https://doi.org/10.1016/j.ajhg.2008.07.001.

**Supplementary Table 5. Simulation results for verification scenario.**

| Years | Total Population (0.99 CI) | Deaf individuals (0.99 CI) | Allele frequency (0.99 CI) | Genotype frequency (0.99 CI) |
|---|---|---|---|---|
| 0 | 200000.00 (±0) | 40.00 (±0) | 0.0144 (±0) | 0.0002 (±0) |
| 20 | 223605.16 (±0.34) | 81.78 (±0.77) | 0.0143 (±0.00) | 0.0004 (±0.00) |
| 40 | 249804.05 (±1.64) | 54.23 (±0.53) | 0.0140 (±0.00) | 0.0002 (±0.00) |
| 60 | 279543.91 (±24.23) | 68.27 (±0.37) | 0.0138 (±0.00) | 0.0002 (±0.00) |
| 80 | 311873.97 (±5.89) | 75.54 (±0.12) | 0.0136 (±0.00) | 0.0002 (±0.00) |
| 100 | 348273.93 (±33.53) | 84.71 (±0.53) | 0.0135 (±0.00) | 0.0002 (±0.00) |
| 120 | 387955.33 (±96.72) | 125.52 (±1.02) | 0.0134 (±0.00) | 0.0003 (±0.00) |
| 140 | 433359.17 (±26.34) | 116.25 (±1.39) | 0.0131 (±0.00) | 0.0003 (±0.00) |
| 160 | 483468.16 (±53.33) | 116.46 (±0.12) | 0.0130 (±0.00) | 0.0002 (±0.00) |
| 180 | 541026.04 (±28.51) | 119.26 (±0.43) | 0.0128 (±0.00) | 0.0002 (±0.00) |
| 200 | 605149.56 (±106.64) | 127.34 (±0.03) | 0.0126 (±0.00) | 0.0002 (±0.00) |
| 220 | 675782.83 (±35.08) | 135.03 (±0.15) | 0.0124 (±0.00) | 0.0002 (±0.00) |
| 240 | 754837.21 (±8.93) | 188.29 (±3.01) | 0.0123 (±0.00) | 0.0002 (±0.00) |
| 260 | 844837.41 (±33.44) | 216.96 (±0.59) | 0.0122 (±0.00) | 0.0003 (±0.00) |
| 280 | 942631.00 (±174.04) | 288.05 (±0.22) | 0.0121 (±0.00) | 0.0003 (±0.00) |
| 300 | 1053543.33 (±90.37) | 344.49 (±0.40) | 0.0120 (±0.00) | 0.0003 (±0.00) |
| 320 | 1177318.19 (±101.03) | 430.05 (±5.30) | 0.0119 (±0.00) | 0.0004 (±0.00) |
| 340 | 1315716.74 (±226.88) | 508.70 (±8.00) | 0.0118 (±0.00) | 0.0004 (±0.00) |
| 360 | 1472053.45 (±34.74) | 607.71 (±4.68) | 0.0117 (±0.00) | 0.0004 (±0.00) |
| 380 | 1646120.66 (±554.41) | 704.94 (±4.03) | 0.0116 (±0.00) | 0.0004 (±0.00) |

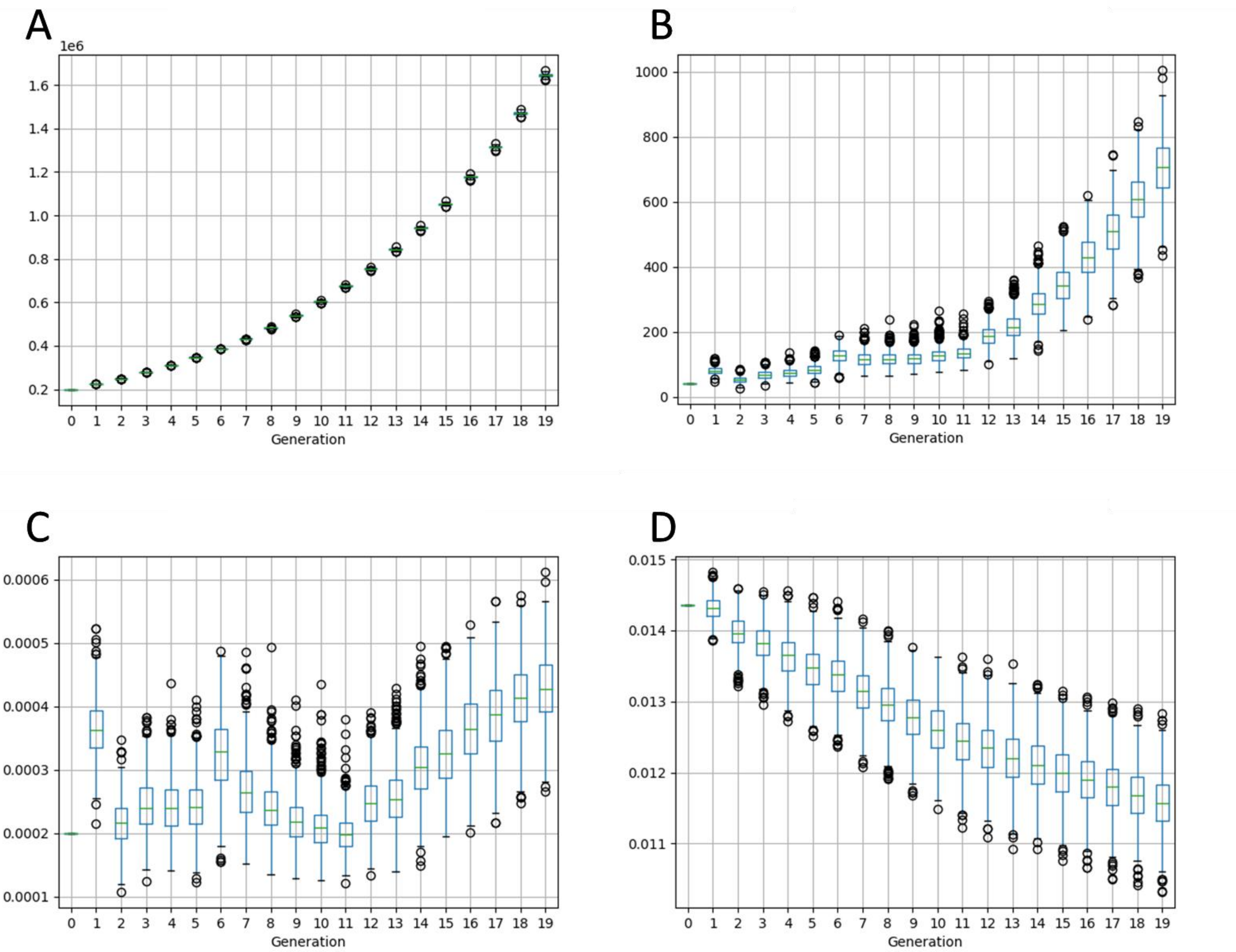


**Supplementary Figure 6.** Simulation results for verification scenario. **A** – Total population size. **B** – The number of deaf individuals. **C** – Proportion of recessive mutant homozygotes in total population. **D** – Frequency of recessive mutant allele in total population
